# The quantified baby: real-world use of infant sleep monitoring technologies and its impact on parent mental health and medical decision-making

**DOI:** 10.1101/2025.05.26.25328367

**Authors:** Jill A. Dosso, Katelyn A. Teng, Katherine T. Roy, Julie M. Robillard

**Author notes:** **Correspondence:** Julie M. Robillard, PhD;, BC Children’s and Women’s Hospitals B402 Shaughnessy, 4480 Oak St. Vancouver, BC, V6H 3N1, Canada.

## Abstract

**Background:** Managing sleep is a challenging experience in early parenthood, and infant sleep problems are associated with negative outcomes within the family. A large market of devices to monitor infants’ real-time health information during sleep has emerged, including smart cameras, under-mattress sensors, and wearable devices. The impacts of these products on maternal and parental mental health and medical decision-making are poorly understood.

**Methods:** We performed a systematic search for products detecting health data from sleeping children on the global retail platform Amazon in March 2023. A total of 11,262 unique reviews from 48 eligible products were retrieved from the US, Canada, UK, and Australia sites and subjected to sentiment and thematic analyses to capture the characteristics of user families, contexts of device use, and impacts on maternal and child health.

**Results:** Parental anxiety and infants’ high-risk medical status were cited as main reasons to purchase products. When devices worked well, their use was associated with improved parental sleep quality and decreased anxiety. However, poor device performance was commonly reported and was linked to increased parental stress and anxiety and disrupted child sleep. Users reported making medical decisions based on device output. Price, privacy, and unsafe use of devices emerged as ethical issues.

**Conclusions:** Use of a smart sleep device in the home is common and has implications for the health of both children and adults. Benefits and harms must be understood by parents and healthcare providers in order to support evidence-based decision-making around their use.

**Key Messages:** *What is already known:*

- Many households monitor sleeping children’s health using emerging “smart” commercial products such as cameras, sensors, and wearables.
- The medical community is not yet prepared to offer evidence-based recommendations about these practices, despite their widespread adoption and potential intersection with the healthcare system.

*What this study adds:*

- By conducting a content analysis of a large corpus of consumer reviews, we find evidence that these devices impact maternal and parental mental health, children’s sleep quality and safety, and family usage of healthcare services.

*How this study might affect research, practice or policy:*

- These results will inform future development of recommendations for sleep device use.
- With a better understanding of how these devices affect user families, parents can be more equipped to make evidence-informed decisions for their family’s health.

## Introduction

Managing household sleep can be a challenging early parenthood experience. Severe problems with infant sleep are associated with negative outcomes, including elevated levels of postpartum depression, postpartum anxiety, maternal anger, and intimate partner violence (Cook et al., 2020; Ou et al., 2022). Insufficient parent sleep is linked to accelerated biological aging and future disease risk (Carroll et al., 2021). Almost all parents wish to change something about their child’s sleep (Mindell et al., 2020).

A large market of devices ($1B USD per year worldwide) intended to support and monitor baby sleep has emerged (Frankel, 2021; Willick, 2022). While baby movement monitors have been available since the 1990s (Angelcare, 2020), commercial products that track infants’ biological data during sleep have rapidly increased in popularity in the last ten years. Early adopters of smart sleep devices were disproportionately highly educated first-time parents, and many reported “peace of mind” and improved adult sleep as initial reasons for purchasing (Dangerfield et al., 2017). Users may be motivated to use child smart sleep devices to prevent sudden infant death syndrome (SIDS) though companies are generally careful to avoid explicit claims to that effect (Pretorius et al., 2020; Willick, 2022). The US Food and Drug Administration, the Canadian Ministry of Health, and the Canadian Paediatric Society all do not recommend any baby products that claim to prevent SIDS, and warn that such devices may pose suffocation risks or lead to a false sense of security (Food & Drug Administration, 2019; Minister of Health, 2021).

While clinical tools are used under different circumstances from commercially available sleep monitors, they output similar data. Parents given monitors (such as apnea monitors and pulse oximeters) following a hospital intervention can experience increased depression, hostility, distress, and fatigue, and decreased social quality of life compared to controls (Abendroth et al., 1999; Ferro et al., 2022; Fu & Moon, 2007; Williams et al., 1999). Whether these patterns are mirrored by families voluntarily using sleep monitoring devices with healthy children is not yet known.

To date, research on the impact of infant sleep health monitoring has largely focused on individual wearable products. Studies have considered technical features of the devices (Bonafide et al., 2017, 2018; Grooby et al., 2023), user values for hypothetical products (Gaunt et al., 2014), consumer purchasing decisions and device acceptance (Hasan & Stannard, 2023), and small-*n* situated studies of families’ experiences (Leder Mackley et al., 2022; Wang et al., 2017). One large manufacturer-led study examined family health impacts and reported of reduced anxiety, better sleep quality, and increased adherence to Safe Sleep guidelines (Dangerfield et al., 2017). However, reductions in touch or in-person checking have also been reported (Leder Mackley et al., 2022; Nelson, 2008; Wang et al., 2017). One study found instances of parents relying on smart sleep monitors to facilitate less-safe sleep practices (Pretorius et al., 2020). Whether these trends hold true for users who ultimately discontinue use, for later product adopters, and for users of other categories of sleep monitoring devices are all open questions.

To address these gaps, the aim of the present work is to characterize real-world home use of commercial health monitors for sleeping children. We focus on its impacts on adult mental health and behaviour and on children’s health. We further differentiate these effects across three monitoring modalities: wearable sensors, environmental sensors, and smart cameras.

## Methods

### Data collection and sampling

We conducted a systematic search of Amazon.com to collect a dataset of infant sleep monitoring devices on March 13-16, 2023. Search terms were selected based on iterative test searches, with the final search strategy including a combination of “baby” with each of the following terms: “sleep,” “night,” “wake,” “alarm,” “safe,” and “smart.” The first 300 listings returned for each combination of terms were reviewed, as pilot testing revealed that all relevant listings appeared within this set. In cases where a single listing contained multiple versions of a product (e.g., different colours or generations), all were captured.

To prevent search returns from being tailored by online history or geographic location, the search was conducted using a browser in Incognito mode, with the Privacy Badger and uBlock Origin extensions activated. Privacy Badger blocks third-party trackers, including advertisements that track the user, and prevents them from loading content into the browser. The uBlock Origin extension is a content-blocker that uses preset filter lists to block all third-party network requests. Browsing data were cleared between each search. Amazon.com was set to deliver to a continental US zip code and was not logged in to an Amazon account.

Once all product listings were identified (Table 1), reviews were downloaded from English-speaking Amazon websites including the USA, Canada, the United Kingdom, and Australia using AMZShark software and automatically deduplicated. Product pages corresponding to each product’s Amazon Standard Identification Number (ASIN) were downloaded. As this study did not involve human subjects or patient data, no institutional ethics approval was obtained. Data for this project was captured directly from public contributors to online fora. As this represents a pilot investigation into a novel area of research for the authorship team, no patient/public partnerships were establish prior to data collection to guide the design, reporting or dissemination of this research.

**Table 1.**
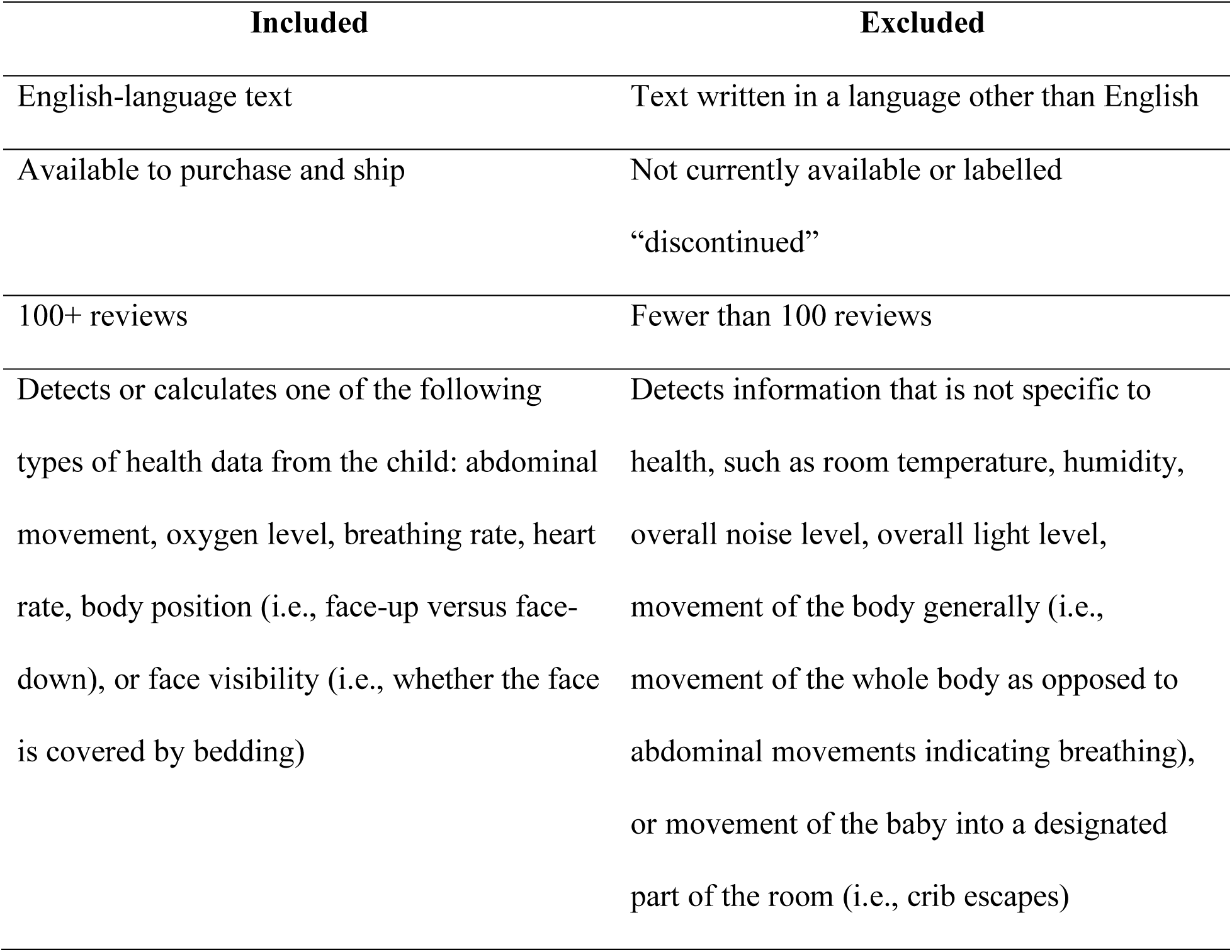
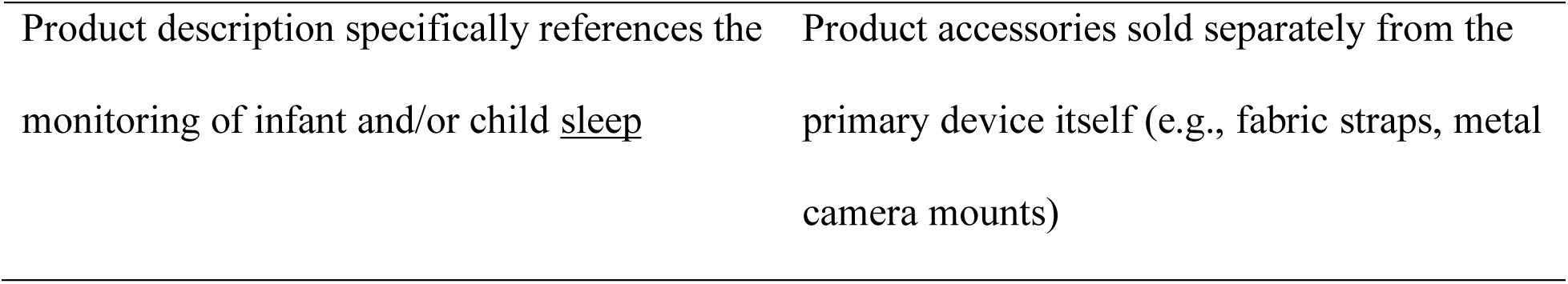
Inclusion and exclusion criteria for product listings.

| Included | Excluded |
| --- | --- |
| English-language text | Text written in a language other than English |
| Available to purchase and ship | Not currently available or labelled<br>“discontinued” |
| 100+ reviews | Fewer than 100 reviews |
| Detects or calculates one of the following types of health data from the child: abdominal movement, oxygen level, breathing rate, heart rate, body position (i.e., face-up versus face-down), or face visibility (i.e., whether the face is covered by bedding) | Detects information that is not specific to health, such as room temperature, humidity, overall noise level, overall light level, movement of the body generally (i.e., movement of the whole body as opposed to abdominal movements indicating breathing), or movement of the baby into a designated part of the room (i.e., crib escapes) |
| Product description specifically references the monitoring of infant and/or child <u>sleep</u> | Product accessories sold separately from the primary device itself (e.g., fabric straps, metal camera mounts) |

### Sentiment analysis

Sentiment analysis was conducted for the full dataset using the tidytext program and visualized with ggplot2 in R (Silge & Robinson, 2017; Wickham, 2016). Reviews were broken down into their constituent words, and emotion and sentiment scores for each word was assigned using the ‘nrc’ lexicon (Mohammad & Turney, 2013). In this lexicon, words are categorized (yes/no) as belonging to the eight emotion categories of anger, anticipation, disgust, fear, joy, sadness, surprise, and trust, as well as two valences (positive/negative). We excluded the names of the products as well as extremely common English words (e.g., “the,” “of”) from our analysis using the tidytext stop_words dataset. The average sentiment of each review was calculated as the number of “positive” words minus the number of “negative” words.

### Content analysis

Reviews were coded following an inductive content analysis approach. Three members of the research team reviewed 100 reviews to create an initial coding guide, identifying themes and subthemes as they emerged from the data without a predetermined framework. A new set of 30 reviews was then coded independently by two researchers, followed by an inter-rater reliability calculation. Disagreements and newly arising code categories were discussed, with corresponding changes to the coding guide. This process was repeated for a new set of 30 reviews until the inter-rater reliability exceeded 85% and both coders endorsed the final coding guide. Then, a new, randomly sampled 10% of all reviews (*n* = 1128, divided evenly across the three product categories) was generated, divided between coders, and coded using the final coding guide.

## Results

### Product characteristics

Our search strategy yielded 26 listing pages containing 48 eligible products (Figure 1): four under-mattress sensor pads (three sold with cameras), 29 wearables (11 diaper clips, 18 foot wraps), and 15 cameras (three sold with a “breathing band” and/or “smart sheet” for calibration). The median price for a listing was $190 USD (range: $60-500). Product listings claimed that products could monitor breathing or abdominal movement (*n* = 16), body position or “rollover” (*n* = 10), provide sleep analytics (*n* = 7), monitor oxygen levels (*n* = 5), and monitor heart rate (*n* = 5).

**Figure 1.**
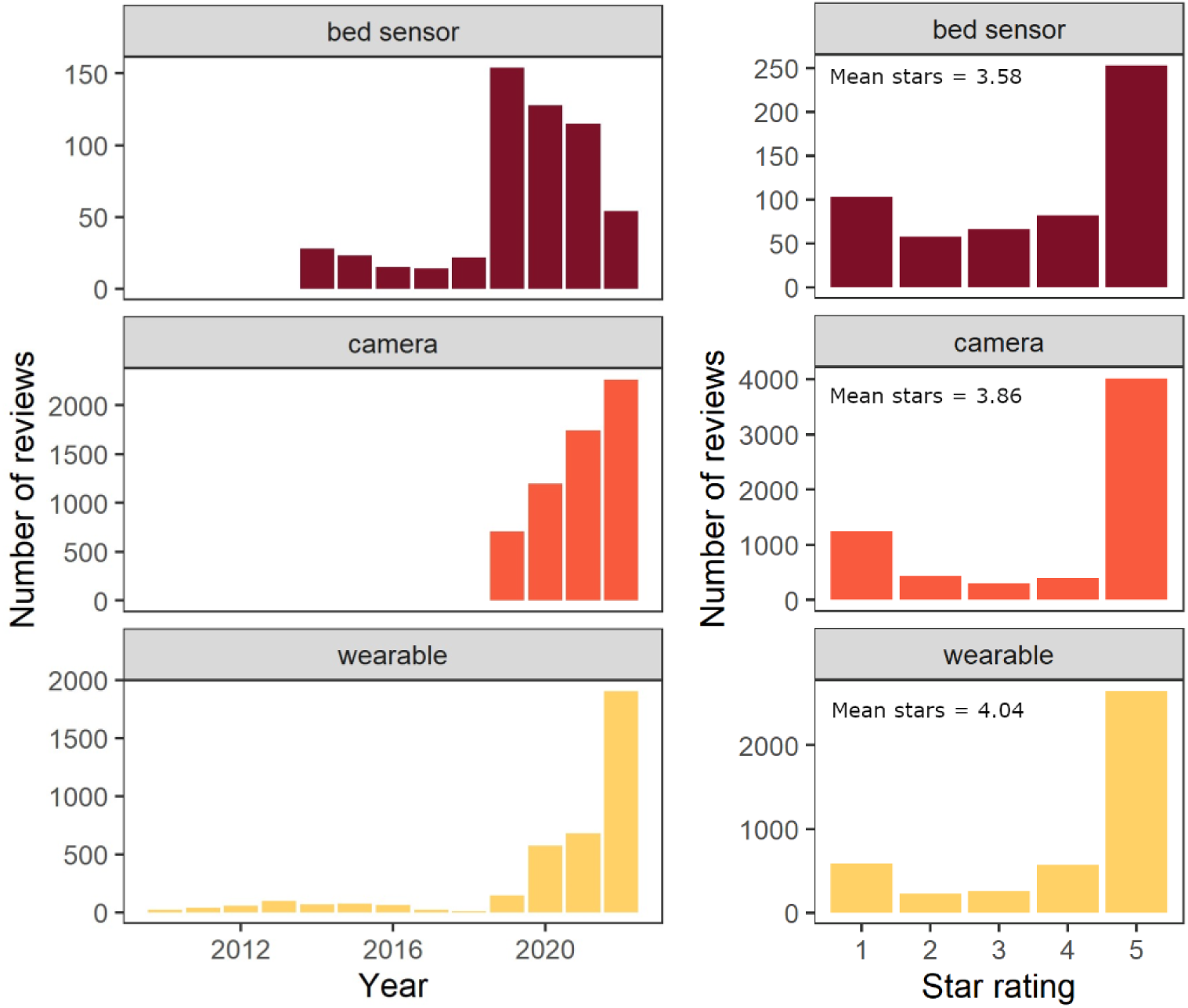
Left panels: Number of reviews recorded for product categories across time, 2010-2022. Note that the y-axis varies across product categories with fewest reviews for bed sensors. Right panels: Distribution of stars across all ratings inclusive of 2023.

There were 11,262 unique reviews following deduplication (*n* = 6414 camera, *n* = 4286 wearable, *n* = 562 bed sensor). Overall, 57% of reviews were 5/5 stars and 20% of reviews were 1/5 stars. Reviews had a mean of 81 words (range 1–1162) and 71% were based on verified purchases. We generated authenticity “Report Cards” (reviewmeta.com) for the products whose proportion of verified reviews was below this average, and none received a score of “Fail.”

### Sentiment within reviews

Reviews typically contained more positive than negative words (range: -11 to +40, mean +2.86), with camera reviews scoring most positively (mean +3.25, standard error 0.05) followed by bed sensors (+2.90, 0.20). Wearable reviews were the least positive (+2.27, 0.05). Reviews predominantly contained words associated with positive emotions such as joy, anticipation, and trust including “peace,” “love,” “recommend,” and “baby” (Figure 2). Frequent words like “money,” “alarm,” “alert,” “battery,” and “diaper” point were associated with anger, disgust, and fear. There was a significant effect of product type on review sentiment as revealed by a one-way ANOVA (*F*(2, 10331) = 89.3, *p* <.001). Specifically, wearable reviews were more negative than the other two categories (both *pbonf* < .001), while bed sensors and cameras did not differ (*pbonf* = .098).

**Figure 2.**
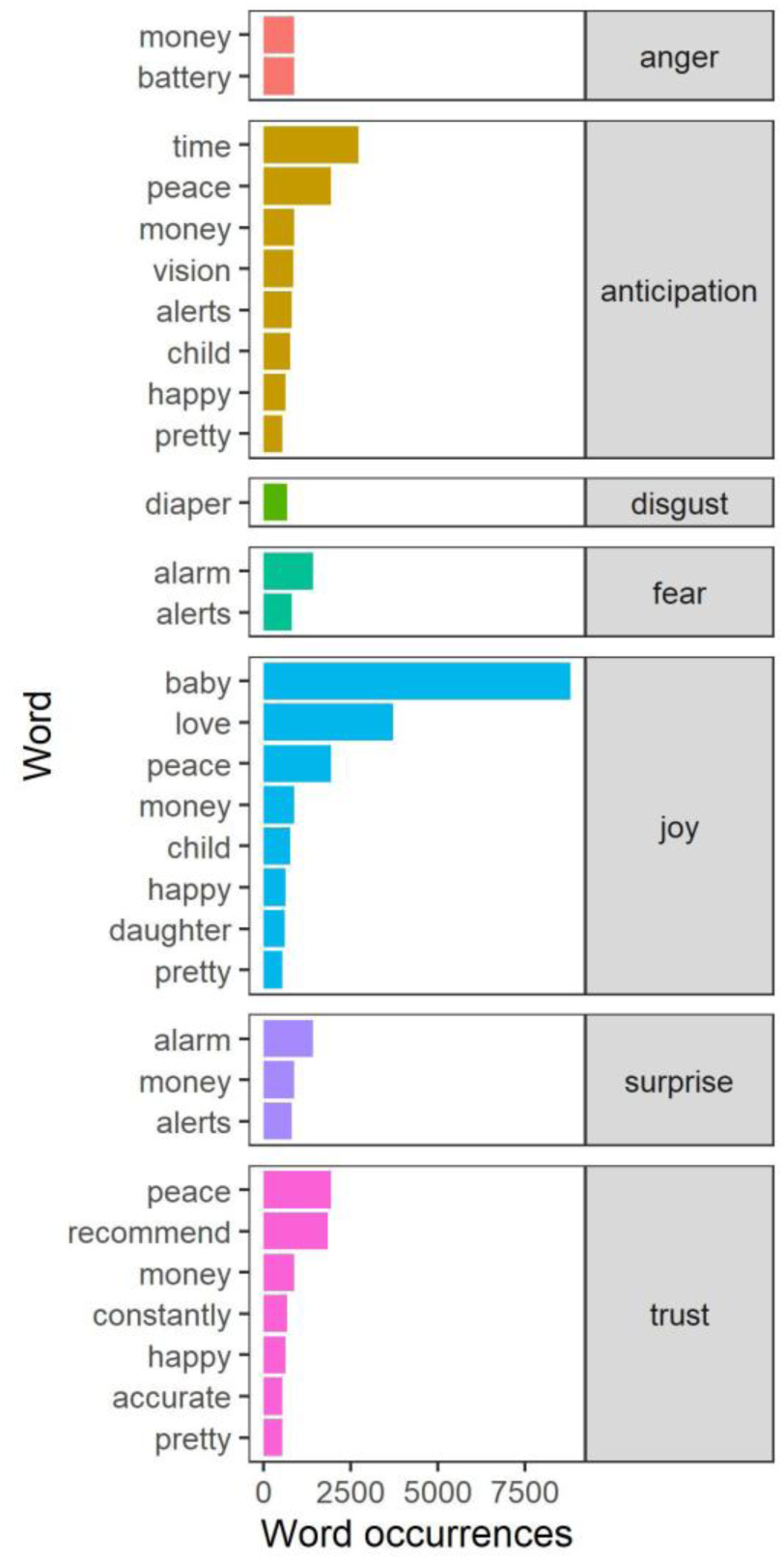
Words occurring in at least 500 total reviews across products and corresponding to the emotions of the ‘nrc’ lexicon (no words met this threshold for the emotion category of “sadness.”)

### Content analysis of reviews

Reviews were 3.4 times more likely to make positive statements about the products as a whole (e.g., “great camera”; 56.8%; *n* = 641) than negative statements (e.g., “so disappointed”; 16.5%; *n* = 186).

### Family description

Review-writers made statements about their own mental health status as it related to device purchase (7.0%; *n* = 79; Figure 3). The majority related to fear and anxiety, using language like “paranoid,” “neurotic,” or “worry-wart,” and describing worries around leaving the baby alone. Some referenced clinical diagnoses like postpartum anxiety or generalized anxiety disorder. Many explicitly named fears about SIDS or breathing problems.

**Figure 3.**
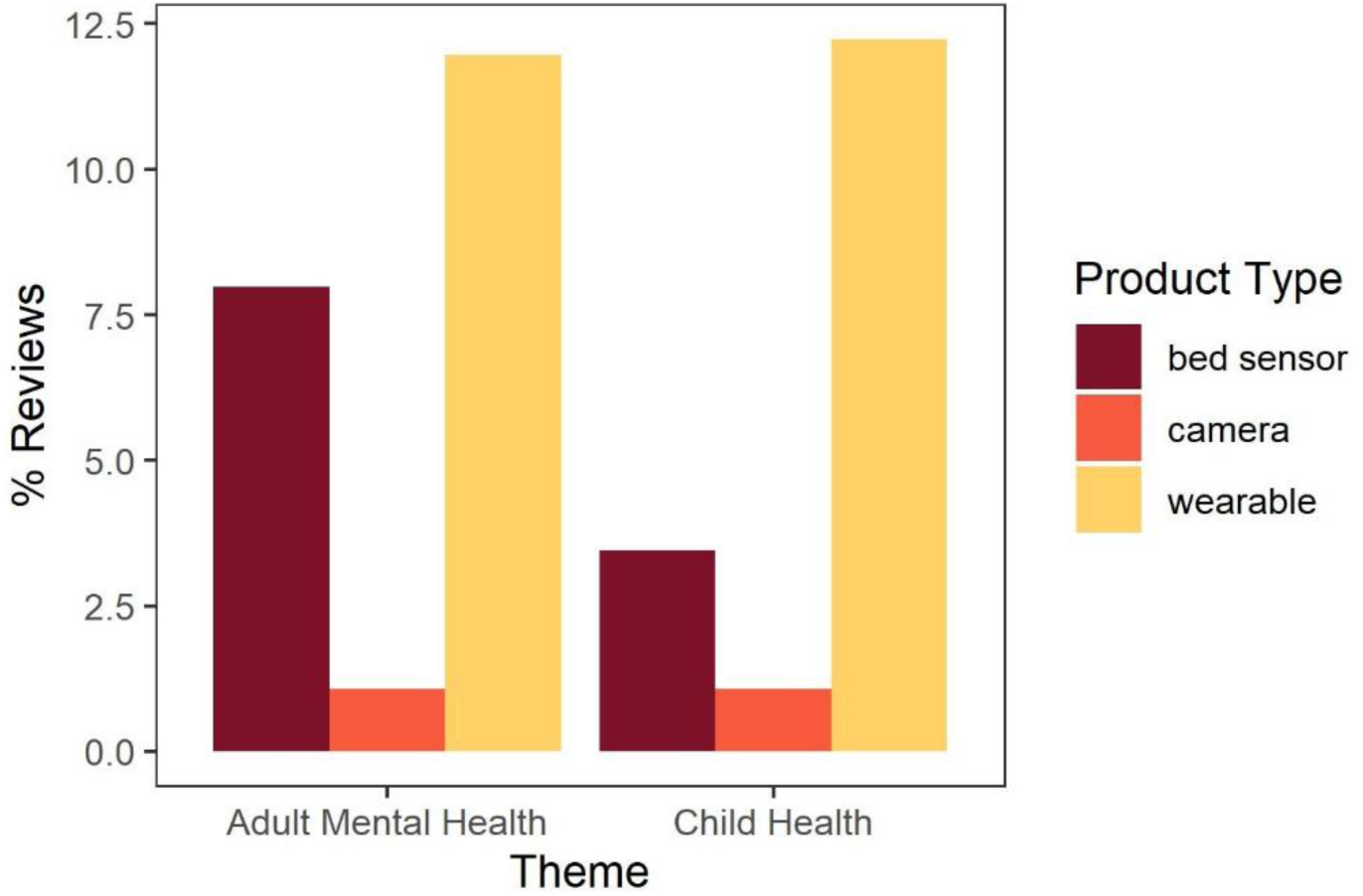
Frequency of references to the mental health status of adult family members and the medical history of young children in the context of device purchase or use, particularly when reviewing wearable products.

Some reviews also described children’s health status or health history (5.6%; *n* = 63 total; Figure 3).The most common conditions that were mentioned were prematurity or NICU stay, breathing issues and asthma, bouts of sickness including colds, congestion, and RSV, and reflux. This theme was most common among reviewers of wearables. A small number of reviews described a child’s general temperament as wiggly, fussy, or easily upset (0.5%; *n* = 6).

Children’s age during use was referenced (9.0%; *n* = 102 reviews, with some reviews mentioning multiple children). Product use was described for premature infants, newborns, older babies, and toddlers. Many reviewers reported using devices from birth onward.

### Context of device use

Many reviews (15.7%; *n* = 177) specified the location of the sleeping child during use. Most referred to children sleeping alone in cribs or bassinets, but a small number of wearable users (Table 1) referenced co-sleeping, car-seats, strollers, and a baby swing. When parents referenced their own location during product use (5.9%; *n* = 67), they were typically in other parts of the same house, though some mentioned bringing devices along for travel, or checking children while under the care of others (e.g., “they’re home with dad.”)

Infant body position was mentioned (3.5%) by bed sensor (*n* = 14) and wearable (*n* = 26) reviewers. Devices reassured parents when children began side-sleeping or sleeping prone as part of typical development. A few described using the products (two wearables and one bed sensor) to facilitate placing very young children in prone positions to sleep, e.g., “Our little one would not sleep on his back from day one. After a week and a half we put him on his tummy and he slept like an angel, however I didn’t sleep a wink. […] I was finally able to get some sleep when our dear [product] came.” Writers also referenced the role of these products during development transitions (2.0%; *n* = 22) such as learning to roll and moving to a new bed.

### Impact on adult behaviour

Adults reported modifying their own behaviour as a function of device output. Specifically, they physically checked on children less often (*n* = 14, e.g., “Gave me peace of mind that I wouldn’t have to repeatedly check her at night!”), adjusted children’s sleeping environments (*n* = 8) and made scheduling decisions (*n* = 7). A few referenced using cameras in modifying child sleep (*n* = 3).

### Impact on adults

When devices worked as intended, the most common mental health impact for adult users was reduced anxiety or “peace of mind” (*n* = 261; 23%; Figure 4), followed by improved adult sleep (10.5%; *n* = 119), and these were often co-reported (e.g., “It gives me peace of mind which means I get more sleep at night”). Two reviews reported increased anxiety when devices were functioning properly.

**Figure 4.**
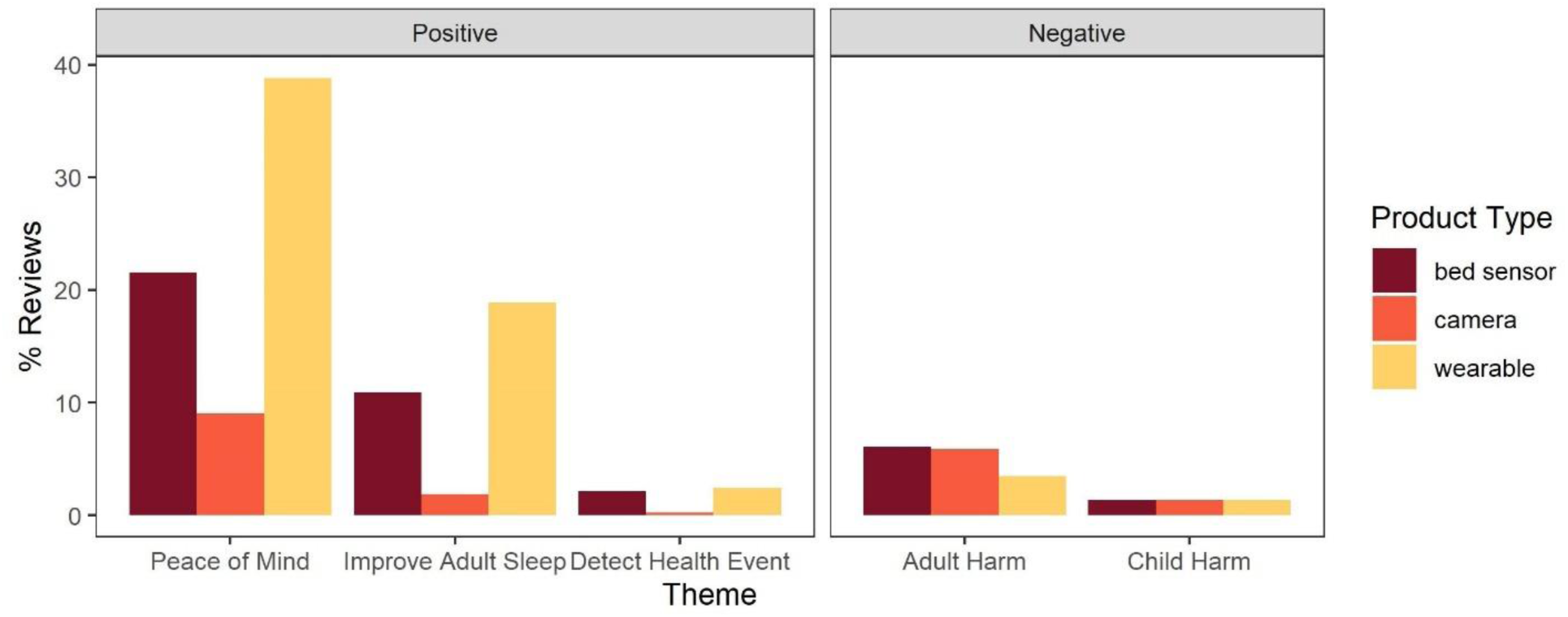
Reviewer-reported impacts of sleep devices on adults and children. Health interventions were performed in response to breathing-related alarms. Adult harms were stress, fear, and awakenings, while child harms were predominantly unnecessary awakenings. Positive impacts took place when devices worked properly while negative impacts took place when devices did not.

### Impact on children

Child health events or interventions were documented in the context of devices working properly (2.0%; *n* = 23 reviews; Figure 4). Many of these described instances of products detecting apparent breathing problems (Table 3). In nine instances, parents took medical actions based on device alarms (*n* = 5 wearable, *n* = 4 bed sensor). Specifically, three describe rousing unresponsive infants, three describe taking ill children to emergency services, and three describe delivering home health interventions (Table 3).

**Table 3.**
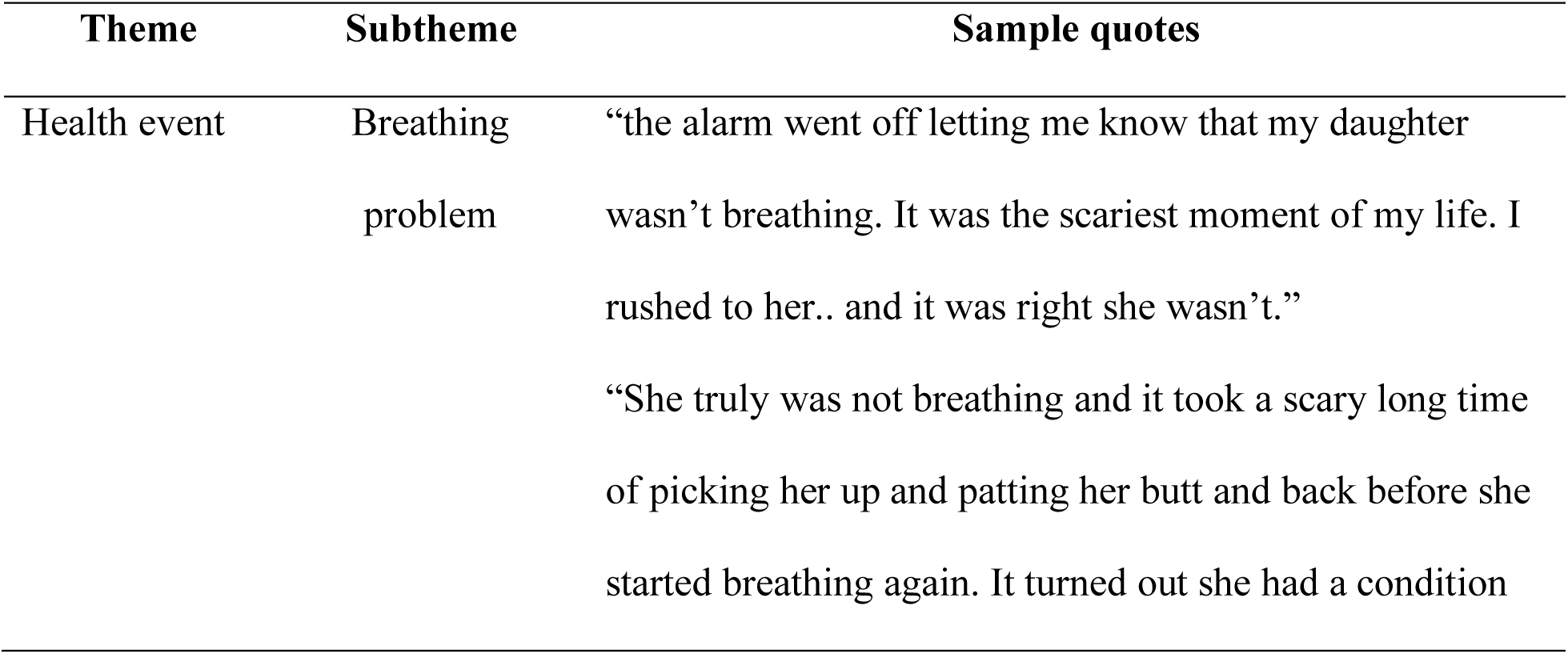
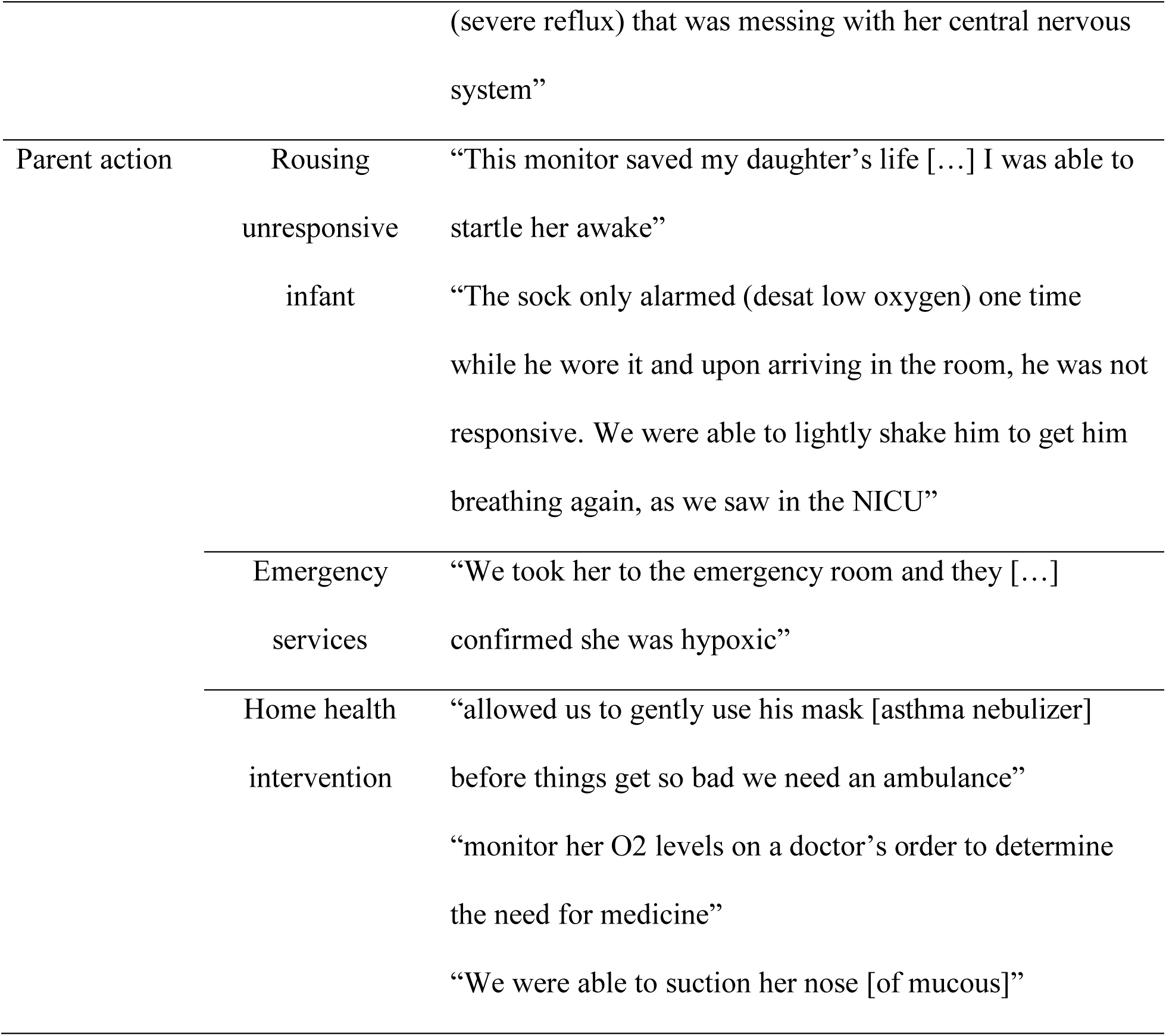
Review content describing children’s health events and parental actions taken.

| Theme | Subtheme | Sample quotes |
| --- | --- | --- |
| Health event | Breathing problem | “the alarm went off letting me know that my daughter wasn’t breathing. It was the scariest moment of my life. I rushed to her.. and it was right she wasn’t.”<br>“She truly was not breathing and it took a scary long time of picking her up and patting her butt and back before she started breathing again. It turned out she had a condition |
|  |  | (severe reflux) that was messing with her central nervous system” |
| Parent action | Rousing<br>unresponsive<br>infant | “This monitor saved my daughter’s life [...] I was able to startle her awake”<br>“The sock only alarmed (desat low oxygen) one time while he wore it and upon arriving in the room, he was not responsive. We were able to lightly shake him to get him breathing again, as we saw in the NICU” |
|  | Emergency<br>services | “We took her to the emergency room and they [...] confirmed she was hypoxic” |
|  | Home health<br>intervention | “allowed us to gently use his mask [asthma nebulizer] before things get so bad we need an ambulance”<br>“monitor her O2 levels on a doctor’s order to determine the need for medicine”<br>“We were able to suction her nose [of mucous]” |

With respect to non-medical impacts on children, six reviews described soothing effects of bed sensors and cameras on children through two-way talk features, music, and sounds, and five describing devices waking children as part of their normal functioning such as flashing bright lights.

### Impacts of poor device performance

Nearly one-third of all coded reviews (32.2%; *n* = 363) described some type of poor device performance, and one-fifth of these (6.9%; *n* = 78) indicated that had stopped using the device due to these issues or planned to stop. Performance issues primarily negatively impacted children in the form of unnecessary wakings, either by alarms or unnecessary parent checks (Figure 4; 1.6%; *n* = 18). One review stated that a child had been left crying due to malfunction, and another described a camera that dangerously overheated. Adults themselves also experienced negative impacts when devices malfunctioned (5.0%; *n* = 56; Table 4; Figure 4). These included reports of being stressed, frustrated, afraid, woken, or kept awake.

**Table 4.**
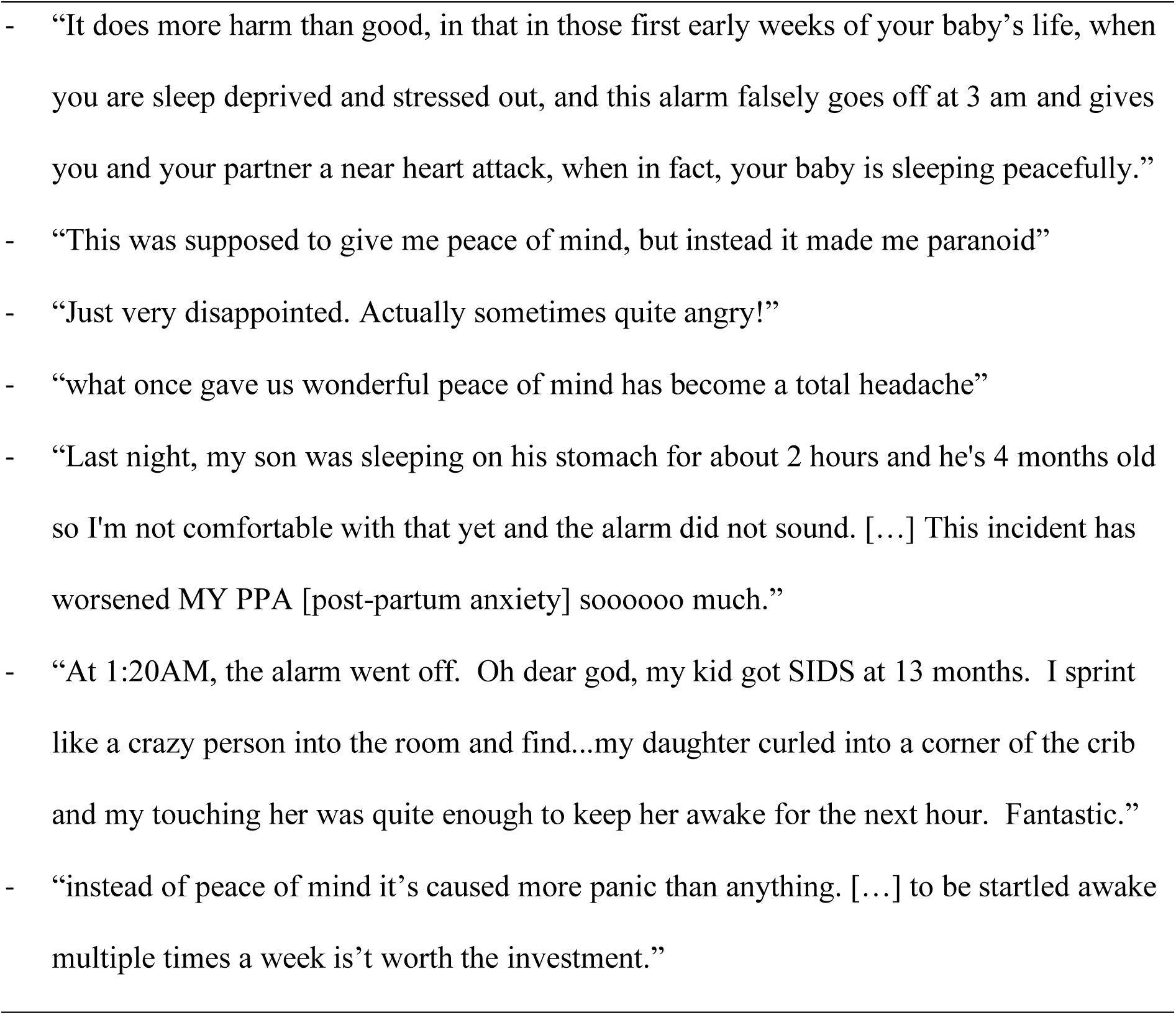
Negative impacts of poor device performance for adult family members.

### Ethical considerations

The most common ethical theme that emerged in coding was price and access (21.4%; *n* = 241). Price was sometimes referred to as high but worth it (e.g., “If you can afford it, get it!”), though other felt that prices were too high for the value being delivered (“just not worth it and for the price there are better monitors”). Price as a barrier to accessing high-quality devices was also mentioned within this theme (“after saving for months (because we’re low-income) and finally purchasing this camera, I’m pretty upset. I feel swindled.”) Privacy concerns such as hacking were also referenced (0.5%; *n* = 6; “it is a Wifi monitor and there for is susceptible to hacking”).

## Discussion

While sensors to monitor infant breathing have a long history of use in clinical contexts, their widespread commercial availability and the normalization of their use for healthy children are more recent phenomena. As the interface through which millions of parents monitor their children daily, these devices have enormous potential to impact maternal mental health, decision-making on behalf of children, and use of healthcare resources.

Our dataset consists of over eleven thousand product reviews containing personal, emotional details about families’ sleep challenges, household finances, and medical histories. This data has high ecological validity as user-generated online content is a rich source of data about parents’ experiences and decision-making related to children’s health (Pretorius et al., 2020; Tougas et al., 2018). We verified the quality of our dataset using a third-party tool and the majority of all reviews were associated with registered purchases. All three independent coders who handled the reviews (totaling hundreds of reviews per coder) reported that they were relevant to the correct products and did not show evidence of manipulation, providing us with a high degree of confidence in their veracity. The data were analysed in two complementary ways. Sentiment analysis was also applied to the full dataset, revealing its emotional content. Targeted, fine-grained thematic content analysis was performed on a subset of reviews to elicit their key themes, an established method to evaluate online health content (Dosso et al., 2023; Feng et al., 2021; Hasan & Stannard, 2023; Pretorius et al., 2020; Tougas et al., 2018).

Understanding the user base for these products is key to creating future public health messaging in this space. In our data, users of these products were largely the parents of children under the age of six months old, and referenced a mental health history characterized by anxiety and worry, with or without a clinical diagnosis. Children’s medical histories, such as prematurity or neonatal hospitalization, were referenced. If healthcare providers would like to counsel families around the purchase of smart sleep monitors, this should be done prenatally. Parents with a history of anxiety and families at a high risk of premature birth or neonatal medical complexity would be particularly relevant populations to receive this information. Discharge from NICU could be another key opportunity for these discussions.

A strength of the present work is our ability to capture the experiences of users who have discontinued using their sleep sensors, unlike the majority of studies which have focused on active users. These missing perspectives are important to capture the true balance of benefits and harms. Reports of poor device performance were very common (one-third of all reviews) and were associated with parental frustration and anxiety, sleep disruption for both parents and children, and discontinuation of product use. For medical staff, nonactionable alarms increase response time to actionable alarms (Bonafide et al., 2015); there is evidence of a similar pattern here, with parents reporting that poor device performance led them to abandon their devices despite wishing for the promised safety functionality. These outcomes, combined with the high price point of many products in this category, raise questions around the ethics of normalizing or recommending the use of these products by all families, especially when equitable access to healthcare is challenging.

In our analysis, we were particularly interested in the impact of smart sleep sensors on parental mental health. The literature contains instances of parents reporting increased mental illness symptoms, describing their checking of device outputs as “obsessive” or “addictive” (Leder Mackley et al., 2022; Wang et al., 2017; Willick, 2022). Some researchers argue that monitors allow for continuous surveillance of the baby which normalizes parental anxiety and creates an unrealistic expectation that parents (particularly mothers) must be always attentive to eliminate all risk of harm (Nelson, 2008). Others raise the possibility that sleep sensors might disrupt the parent-child connection or the formation of parental intuition about children’s needs (Gaunt et al., 2014; Leder Mackley et al., 2022). However, the reverse effect, that sleep monitoring devices reduce parental anxiety, has also been reported (Dangerfield et al., 2017; Leder Mackley et al., 2022). Our data are more consistent with the latter. Parents typically reported a reduction in anxiety or “peace of mind” in association with their use, as well as improved sleep (so long as devices functioned normally). We are not able to discriminate between two scenarios here: in one, anxious families may be justifiably reassured that their infants are well; in the other, families may be inappropriately reassured that devices can be relied on to catch dangerous incidents and may reduce checking or rely on devices during unsafe sleep practices.

A small number of parents reported taking medical actions based on sensor output, including rousing unresponsive infants and seeking out medical services. These outcomes represent the best-case scenario for these products and were more common for wearables and bed sensors than smart cameras. We are not able to objectively determine the nature of these parent-reported medical events. It may be most appropriate to consider events where sleep sensors go off to be “apparent life-threatening events” (ALTEs), a category that carries risk but is not synonymous with SIDS (Fu & Moon, 2007; Sahewalla et al., 2016). A more minor health outcome that also appeared in the data was children being woken by malfunctioning devices or by parents responding to known false alarms. Devices also impact children indirectly by shaping their parents’ decision-making. Some describe checking on their children less often when using sensors. Others used the devices to support children’s sleep outside of typically recommended contexts, including bed-sharing, using swings, and placing young infants in prone positions. We did not find evidence of emotional disconnection between parent and child. Overall, we find strong evidence that children’s health is impacted by the use of these technologies.

Our methodology captures rich data about the benefits and harms of smart sleep device use by a large sample of real families in their communities. However, relying on user-generated content has trade-offs. First, it is likely that certain perspectives are over-represented; families with particularly positive or negative experiences may be more likely to leave product reviews. Second, demographics such as age, gender, location, and socio-economic status are unknown. Third, relying on product reviews cannot capture the dynamics of using these products over time. Further studies can mitigate these issues using experimental and longitudinal methods.

To conclude, in an analysis of over eleven thousand product reviews, we investigated the impact of the widespread use of infant sleep sensors in naturalistic home environments on maternal and child health. Families using smart sleep devices tended to report parental peace of mind and improved sleep. A small minority report that the products detected important medical events, while others used the tools to support unsafe sleep practices. Poor device performance was both common and highly stressful. Wearables and bed sensors were more likely than smart cameras to be purchased by families reporting a pattern of anxiety for parents and medical risk (prematurity and/or NICU stay) for children. Price was an emerging ethical issue. This work is a first step towards the development of evidence-based resources for health experts and parents about real-world risks and benefits of using these devices.

## Declarations

### Funding

This work was supported by a Michael Smith Health Research BC Fellowship to JAD, a Natural Sciences and Engineering Research Council (NSERC) Fellowship to JAD, funding from BC Children’s Hospital Research Institute to JMR and from the BC Children’s Hospital Foundation to JMR.

### Conflicts of Interest

The authors declare that they have no conflict of interest.

### Ethics approval

Not applicable – no human subjects or patient data.

### Consent to participate

Not applicable – no human subjects or patient data.

### Consent for publication

Not applicable – no human subjects or patient data.

### Availability of data and material

The data underlying this study are available by request.

### Code availability

The analysis code supporting this study is available by request.

### Authors’ contributions

JAD: conceptualization, methodology, software, validation, formal analysis, investigation, data curation, writing – original draft, writing – review & editing, visualization, supervision, project administration, funding acquisition; KAT: validation, formal analysis, investigation, data curation, KTR: validation, formal analysis, investigation, data curation; JMR: conceptualization, methodology, writing – review & editing, supervision, project administration, funding acquisition.

## Acknowledgements

This work was supported by a Michael Smith Health Research BC Fellowship to JAD, a Natural Sciences and Engineering Research Council (NSERC) Fellowship to JAD, funding from BC Children’s Hospital Research Institute to JMR and from the BC Children’s Hospital Foundation to JMR. The authors thank Jaya N. Kailley for contributing additional data checking.

## Notes

### Competing Interest Statement

The authors have declared no competing interest.

